# A robust gene expression signature to predict proteasome inhibitor benefit in Multiple Myeloma

**DOI:** 10.1101/2019.12.16.19015024

**Authors:** Joske Ubels, Pieter Sonneveld, Martin H. van Vliet, Jeroen de Ridder

## Abstract

Many cancer drugs only benefit a subset of the patients that receive them, but are often associated with serious side effects. Predictive classification methods that can identify which patients will benefit from a specific treatment are therefore of great clinical utility. We here introduce a novel machine learning method to identify predictive gene expression signatures, based on the idea that patients who received different treatments but exhibit similar expression profiles can be used to model response to the alternative treatment. We use this method to predict proteasome inhibitor benefit in Multiple Myeloma (MM). In a dataset of 910 MM patients we identify a 14-gene expression signature that can successfully predict benefit to the proteasome inhibitor bortezomib, with a hazard ratio of 0.47 (p = 0.04) in class ‘benefit’, while in class ‘no benefit’ the hazard ratio is 0.91 (p = 0.68). Importantly, we observe a similar classification performance (HR class benefit = 0.46, p = 0.04) in an independent patient cohort which was moreover measured on a different platform, demonstrating the robustness of the signature. Moreover, we find that the genes in the discovered signature are essential, as no equivalent signature can be found when they are excluded from the analysis. Multiple genes in the signature are linked to working mechanisms of proteasome inhibitors or MM disease progression. In conclusion, our method allows for identification of gene expression signatures that can aid in treatment decisions for MM patients and provide insight into the biological mechanism behind treatment benefit.

## Background

For many anti-cancer drugs the response varies widely across patients. As many of these drugs are associated with serious side effects, it is essential to identify which drug will maximally benefit the patient. Tools that aid in such decisions, e.g. based on patient-derived genetic or transcriptomic profiles have only been developed for a few treatments and diseases. Most efforts in this direction focus on detecting specific mutations for which it is known that a targeted therapy exists^1^. However, many patients do not carry any mutations that are known to be actionable and in practice only 7% of patients can be matched to a targeted therapy with the highest level of evidence^2^. Moreover, a range of efficacious therapies exist that are non-targeted and can therefore not be matched to a specific mutation. Consequently, there is a clear clinical utility for methods that can more generically predict - at the time of diagnosis - if a patient will benefit from a certain treatment or not.

Multiple myeloma (MM) is characterized by a malignant proliferation of plasma cells, both in the bone marrow and extramedullary sites. MM is considered incurable with a median survival of approximately 6 years^3^. Several driver mutations have been identified in MM^4^, but in most patients no actionable mutations are observed and targeted therapies are therefore not commonly used in MM. Currently, proteasome inhibitors (PIs) are one of the most important components of treatment in MM and since their introduction in the clinic survival has significantly improved^5^. Since the immunoglobulin production of MM cells is higher than healthy plasma cells, they are more reliant on proteasomal degradation of proteins, making them vulnerable to proteasome inhibition^6^. After bortezomib, which was the first PI to be introduced in the clinic for MM, second generation proteasome inhibitors like carfilzomib and ixazomib have recently been approved.

Despite the success of PIs, there is still wide variability in PI response across patients. Substantial efforts have been made to discover what distinguishes responders from non-responders. For instance, several studies have implicated differential expression of genes involved in the unfolded protein response^7^. Other studies describe complex changes in the entire energy metabolism as a potential discriminating factor^8^. Several chromosomal aberrations have also been found to influence bortezomib response, although this effect is still not fully understood ^9,10^. Despite all efforts, there is currently no biomarker capable of determining which patients will benefit from receiving a PI and which would not.

Most of the studies investigating PI response compare gene expression patterns of patients responding well or poor to a certain treatment^7, 11–13^. The identified genes can then be combined into a gene expression signature, which can be applied to classify newly diagnosed patients as good or poor responders. However, a clinically more interesting question is whether a patient will benefit more from a PI than from another treatment. This is a markedly different question than identifying good and poor responders within one homogeneous treatment group. After all, even patients with lower than average survival may still experience benefit from their treatment as their outcome could have been even worse on another treatment. Conversely, a patient with a good survival outcome, could have experienced an equivalently good or even better response on any other treatment. As a result, it is impossible to assign patients to class ‘benefit’ or ‘no benefit’ a priori, since response to another treatment cannot be observed. Therefore, standard methods of identifying gene signatures and classifiers, which rely on the existence of such class labels, are unsuitable for predicting treatment benefit.

In this work we propose a novel method, Simulated Treatment Learning signatures (STLsig), to infer gene signatures that can predict treatment benefit for patients at the moment of diagnosis. We apply STLsig to find a gene expression signature capable of identifying patients for whom treatment with PIs results in better survival than treatment with an alternative treatment. First and foremost, the gene signature should be capable of predicting PI benefit in an independent patient cohort, a feat which has been shown challenging for prognostic classifiers^14^. A second important objective of STLsig is to identify a simple, interpretable model which contains genes that have biological relevance to the molecular mechanism underlying PI efficacy. To enable this, we leverage the core concept of Simulated Treatment Learning (STL), which we proposed previously^15^, that allows training classifiers without having a predefined labelling of patient in e.g. a class ‘benefit’ and ‘no benefit’. However, while our previous method was successful in identifying a model that can predict treatment benefit, these models rely on large numbers of Gene Ontology sets together containing hundreds of genes. This makes interpretation of these models complex. Moreover, many of the gene sets in the model can be removed without harming performance, casting doubt on their importance in the molecular mechanism of PIs.

For these reasons we here propose a different approach which identifies small networks of genes which can be used as a gene signature to predict PI benefit. To obtain a signature for treatment benefit, we aim to form gene networks that combine genes that are complementary in their ability to predict benefit. STLsig achieves this by assessing the predictive ability of all possible pairs of genes and only combining those that achieve a better performance together than when combined with other genes. STLsig is fully data driven and does not rely on any biological knowledge or predefined gene networks as input.

We demonstrate the utility of STLsig on a 910 sample dataset combining three different Phase III clinical trials with MM patients receiving either a treatment with or without the PI bortezomib (from here on referred to as the HTT cohort). STLsig enables discovery of a 14-gene signature that can accurately identify a subset of patients benefiting from bortezomib. We validate this gene expression signature in independent data (the CoMMpass cohort) where we predict benefit for bortezomib or an alternative PI, carfilzomib, demonstrating that the signature is robust and generalizes to other data. Moreover, we established that the signature is unique, meaning that when removed from the dataset no gene expression signature with a similar performance can be found. The genes included in the signature are thus essential for predicting PI benefit. We find that several of the genes in the signature are related to MM or the working mechanisms of PIs. To our knowledge, this is the first approach capable of discovering treatment benefit specific gene signatures without predefined labels.

## Results

### Overview of the algorithm

STLsig relies on the idea that patients exhibiting similar gene expression profiles who received different treatments, can be used to model response to the treatment they did not receive. Similarity between patients needs to be defined by genes relevant to treatment benefit. STLsig therefore derives treatment specific gene networks, which can be used to form a gene expression signature capable of predicting treatment benefit. To train this signature we divide the HTT cohort in a training set (n = 606) and a test set (Fold D, n = 304). The training set is further subdivided into three equal parts, fold A, B and C. We then assess the ability to predict bortezomib benefit for all 5,506,221 gene pairs arising from the genes with a high variance across patients pre-treatment in the HTT training set (n = 3319).

For each patient *j* in fold A, we determine a z-score (zPFS) per gene pair describing the normalized mean survival difference of patient *j* with its most similar neighbours in terms of gene expression that received a different treatment than patient *j*. The survival difference is normalized to obtain a z-score by comparing it to the survival difference found with randomly selected neighbours. A z-score above 0 indicates a larger positive survival difference than expected, a score below 0 a smaller difference. We then test the ability of the gene pair to predict the zPFS score for patients in fold B. We also assess the performance of each gene pair when calculation of zPFS is performed on Fold B and predicted on Fold A. Performance of each gene pair is defined as the mean Spearman rank correlation coefficient between predicted and calculated zPFS values in both folds. A gene pair is retained if it is synergistic, i.e. if the genes in the pair predict zPFS better together than when they are paired with other genes.

We next form a consensus network by repeating the two-fold cross validation five times. Only gene pairs that are found to be synergistic in all repeats and that exceed the median correlation across all gene pairs and all repeats are retained. From this consensus network we extract gene networks, i.e. all connected components.

A gene network can be used to classify patients by using its genes to recalculate a zPFS score for each patient and classifying the top 25% of the patients as class ‘benefit’ and the rest as class ‘no benefit’. To evaluate each gene network, we use this procedure to classify all patients in fold A and B. Subsequently, gene networks are ranked based on the difference between the Cox regression β’s found in class ‘benefit’ and class ‘no benefit’. To build the signature, we sequentially add each network based on this ranking and evaluate the performance of the combined networks on fold C. The different steps of the algorithm are summarized in Figure 1 and explained in detail in the Methods section.

**Figure 1.**
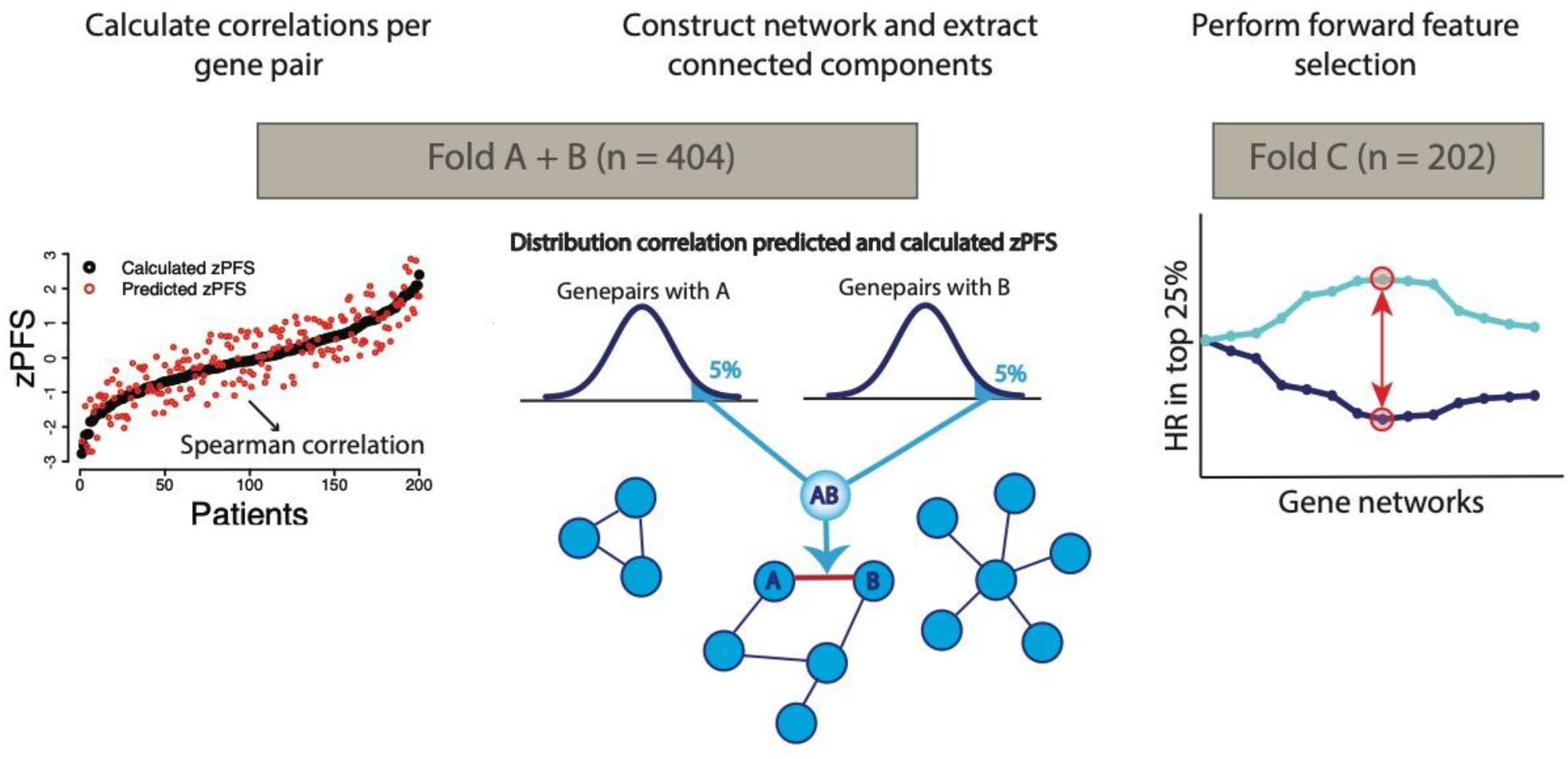
Overview of the construction and selection of the gene networks. First each gene pair is scored on the correlation between predicted and calculated zPFS. Gene networks are then formed by connecting synergistic genes, i.e. genes that are amongst the top 5% partners for each other based on correlation coefficient. The gene networks are then ranked based on difference between Cox regression β in class ‘benefit’ and ‘no benefit’. The signature consists of the combination of gene networks that results in the largest difference in Cox’ regression β between class ‘benefit’ and ‘no benefit’.

### Gene networks yield a 14-gene signature that can predict bortezomib benefit

The consensus network formed as described above contains 617 genes connected by 451 edges and consists of 167 connected components, referred to as gene networks. Of these 167 gene networks, 104 are individual gene pairs. The largest gene network contains 42 genes; the mean number of genes per network is 3.7. We find that the optimal signature is formed by combining the top two ranked gene networks, which are shown in Figure 2. Based on this signature we find a hazard ratio (HR) of 0.49 (p = 0.09, 95% CI 0.22 - 1.11) in class ‘benefit’ (n = 50) and an HR of 0.91 (p = 0.74, 95% CI 0.54 - 1.55) in class ‘no benefit’ (n = 152), on fold C of the HTT cohort.

**Figure 2.**
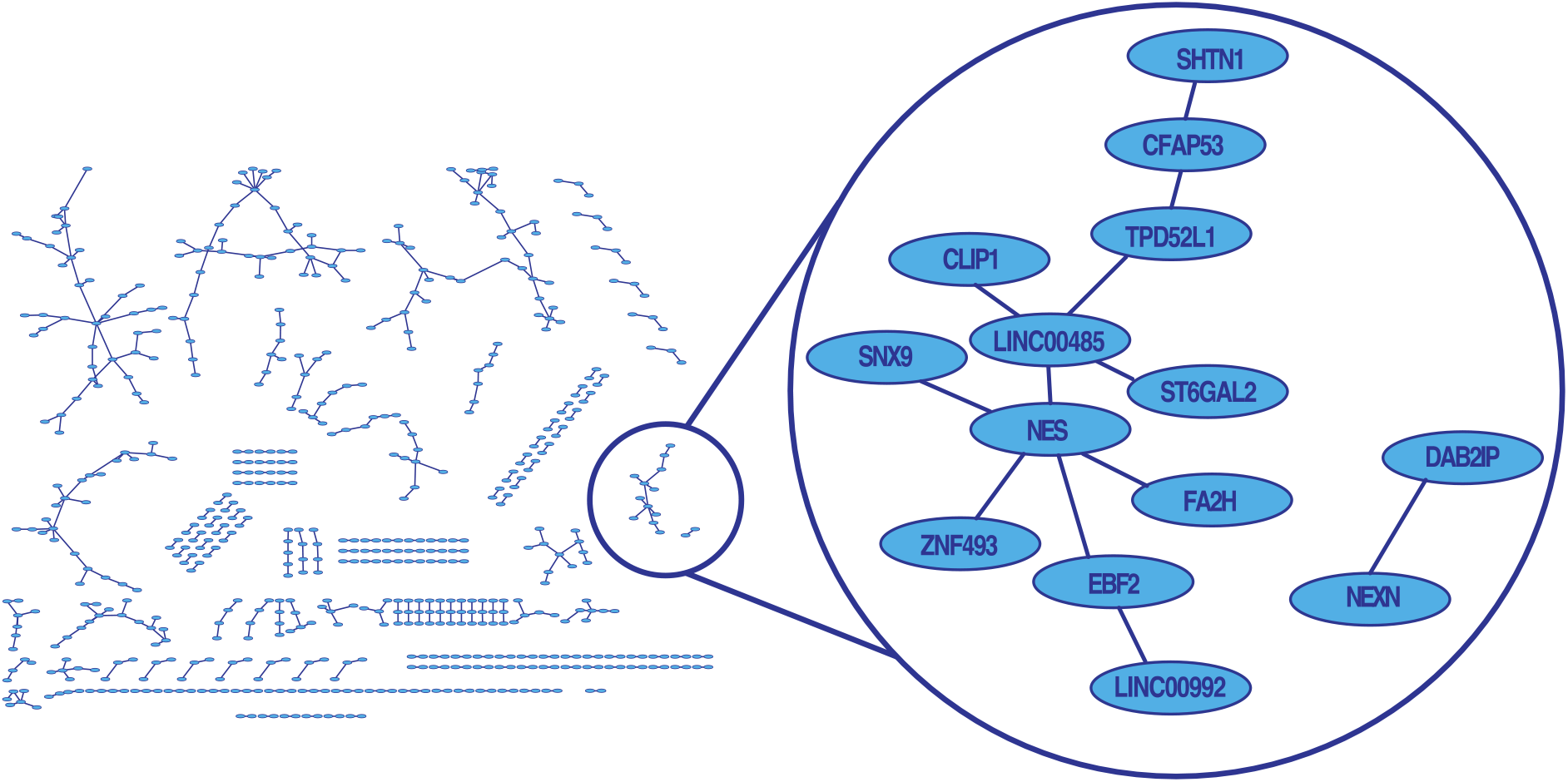
The constructed network with all gene networks. The highlighted networks are those selected by the feature selection procedure and contain the 14 genes in the signature

In clinical deployment of our classifier zPFS scores are not available, as survival of a patient is not known at the moment of diagnosis. In order to assign a zPFS score to a new and unseen patient, we calculate the distance in gene expression space between the new patient and every patient in the training data, which we call the reference set. The predicted zPFS score of the new patient is the weighted sum of the zPFS scores of the patients in the reference set. Weights are determined by the inverse distance, i.e. more similar patients in the reference set contribute most to the predicted zPFS for the new patient (see ‘Methods’ for details). In this manner, we assess the ability of the 14-gene signature to predict benefit in unseen patients, using the 304 patients from the HTT cohort not included in training (Fold D). The HR in favour of bortezomib found in the complete set of 304 patients is 0.75 (p = 0.11, 95% CI 0.53 - 1.06). Figure 3a shows the HR in class benefit obtained using different zPFS thresholds. This shows that a range of thresholds result in an HR below the HR observed in the total dataset, indicating that the predicted zPFS is associated with bortezomib benefit. The optimal class ‘benefit’, i.e. the class ‘benefit’ formed when we select the threshold associated with the lowest HR, comprises 30.6% of the patients which corresponds to a zPFS threshold of 0.326. Using this threshold to define class ‘benefit’ we find an HR of 0.47 (p = 0.04, 95% CI 0.23 - 0.96) in class ‘benefit’ and an HR of 0.91 (p = 0.68, 95% CI 0.60 - 1.39) in class ‘no benefit’ (figure 3b). This establishes that our signature can predict bortezomib benefit in unseen data from the same patient cohort, demonstrating that the signature can be used prospectively to inform treatment choice. Our results indicate that, despite the fact that nearly all MM patients receive a treatment regimen that includes a PI^5^, approximately 70% of patients do not see benefit.

**Figure 3.**
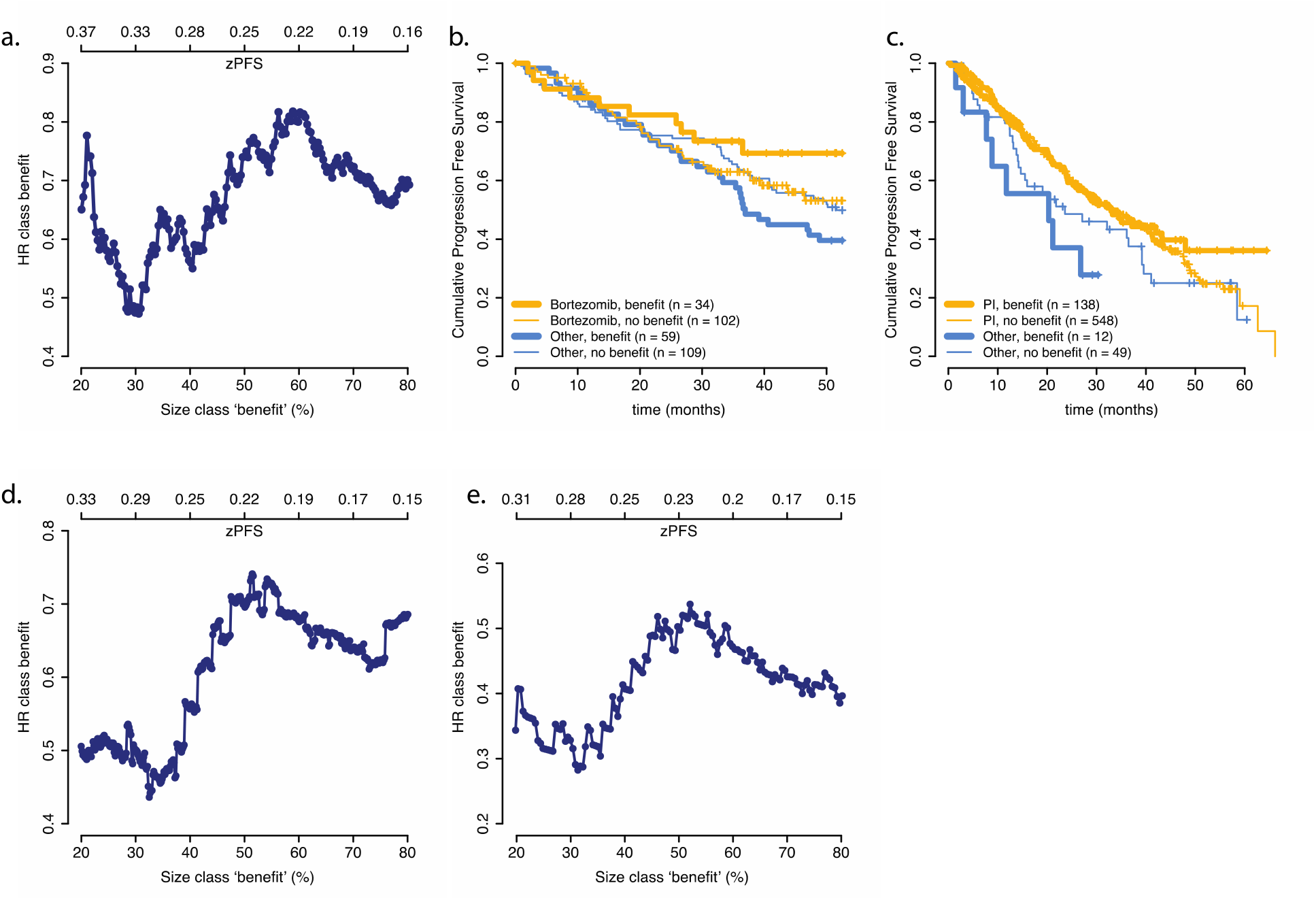
a. HR found in class ‘benefit’ using different zPFS thresholds on the hold out data. b. KM of bortezomib benefit prediction in the hold out data using the optimal zPFS threshold. c. KM of PI benefit prediction on CoMMpass using the optimal zPFS threshold from the hold out data. d. HR found in class ‘benefit’ for bortezomib in CoMMpass, using different zPFS thresholds. e. HR found in class ‘benefit’ for carfilzomib in CoMMpass, using different zPFS thresholds.

### The 14-gene signature achieves robust prediction performance in an independent patient cohort

Gene expression signatures often suffer from cohort-specific fitting and cross-validation within one dataset can thus lead to an overestimation of performance^16^. To obtain a more robust estimate of performance it is essential to perform validation on an external and completely independent cohort. Therefore, to assess the robustness of our signature, we validate its performance in the CoMMpass trial (NCT145429), which represents a completely independent patient cohort which was moreover profiled on a different platform (RNAseq). In contrast to the HTT dataset, which is a randomized clinical trial, the CoMMpass dataset is an observational study. In this study there is no interference with treatment choice and the treatment regimens present in this study thus represent current clinical practice in MM. To bring the CoMMpass RNAseq data in the same space as the microarray reference dataset, we employ a ComBat batch correction (Supplementary Figure 2 and Methods). In the HTT cohort, bortezomib is the only PI used, but In the CoMMpass cohort, two different PIs are used: bortezomib and carfilzomib. To assess the ability of the signature to predict PI benefit, we define a PI treatment arm (n = 686) and a no PI treatment arm (n = 61). When we classify these patients with the 14-gene signature using the threshold optimized on fold D from the HTT cohort, we find an HR of 0.46 (p = 0.04, 95% CI 0.22 - 0.97) in class ‘benefit’ (n = 150) and an HR of 0.79 (p = 0.2, 95% CI 0.55 - 1.13) in class ‘no benefit’ (n = 597) (Figure 3c). Our signature is thus capable of predicting benefit in a completely independent cohort and across platforms, indicating the signal picked up by our classifier is robust and generalizes to the broader MM patient population.

We next assess the performance of the signature for each of the two PIs separately. The majority of the patients in the PI treatment arm received bortezomib (n = 530), while 156 patients received carfilzomib. In the dataset as a whole an HR of 0.75 (p = 0.09, 95% CI 0.54 - 1.04) in favor of bortezomib is found, when compared with patients who did not receive a PI. An HR of 0.42 (p = 0.0004, 95% CI 0.26 - 0.68) is found in favor of carfilzomib. When we evaluate benefit for the bortezomib patients only (excluding patients who received carfilzomib from the analysis), we find an HR of 0.49 (p = 0.06, 95% CI 0.23 - 1.03) in class ‘benefit’ (n = 124) and an HR of 0.84 (p = 0.35, 95% CI 0.58 - 1.21) in class ‘no benefit’ (n = 467). When predicting benefit for the carfilzomib patients only we find an HR of 0.31 (p = 0.06, 95% CI 0.09 - 1.02) in class ‘benefit’ (n = 38) and an HR of 0.45 (p = 0.004, 95% CI 0.26 - 0.77) in class ‘no benefit’ (n =178). These results confirm that our signature is capable of identifying a patient group with more benefit than the population as a whole for each of the PIs separately. Nonetheless it should be noted that the carfilzomib ‘no benefit’ class should rather be considered a ‘less benefit’ class, as a significant HR in favor of treatment with carfilzomib remains, likely due to the highly significant overall carfilzomib HR (0.42). We could not train a carfilzomib specific classifier, as it was absent in the HTT cohort and sample size in CoMMpass is insufficient. Nevertheless, the observation that the bortezomib derived signature can identify a patient group with substantially reduced HRs for carfilzomib treated patients indicates that it is more broadly applicable to PIs in general and not only bortezomib.

We observed that the number of patients classified as ‘benefit’ in the CoMMpass dataset is somewhat lower than on the HTT dataset (20.1% of the patients compared to 30.6% of the patients in fold D of the HTT cohort). When we calculate the HR on the CoMMpass dataset using different zPFS thresholds to define class ‘benefit’, we find that for both bortezomib and carfilzomib the class ‘benefit’ associated with the lowest HR contains approximately 30% of the patients (Figure 3d,e), similar to what was observed in the HTT data. These results show that zPFS score is also associated with benefit at different zPFS thresholds and suggests approximately 30% of MM patients experience more benefit from PI treatment than the population as a whole.

Finally, we evaluated whether our model is specific to proteasome inhibitors and not more generally predictive by testing its performance on lenalidomide, a drug with an immunomodulatory mechanism. In the CoMMpass dataset, 411 patients received lenalidomide and 336 did not (HR of 0.72 in favor of lenalidomide; p = 0.001, 95% CI 0.58 - 0.88). When we define class ‘benefit’ according to the optimized zPFS threshold, 21.0% of patients are stratified into the ‘benefit’ class. We find an HR of 0.80 (p = 0.35, 95% CI 0.51 - 1.27) in class ‘benefit’ (n = 150) and an HR of 0.68 (p = 0.001, 95% CI 0.54 - 0.86) in class ‘no benefit’ (n = 467), hence not improving the HR observed in the lenalidomide dataset as a whole. This clearly shows the signature is predictive specifically for PIs.

### Selected genes and links between them are essential for performance

In the setting of prognostic classification in breast cancer, it is well known that, after removal of the genes in the prognostic signature, a new signature can be found containing a completely different set of genes, that performs equal to the original signature^17^. This phenomenon casts doubt on the value of interpreting the genes in the signature. If biological interpretation is the goal, it is therefore important that no equally performing signature without these genes can be found.

We first investigate the importance of the individual genes in benefit prediction. This is achieved by permuting the expression vector for each gene in the signature 100 times (while the other 13 genes remain unchanged) and using this signature with one permuted gene to predict class ‘benefit’ in fold D of the HTT cohort. Figure 4a shows the mean reduction in HR in class ‘benefit’ after shuffling. These results indicate that each gene is important for the classification performance. The largest effect is observed for DAB2IP, with a mean difference in validation HR of 0.29 (se = 0.06). We also note that correlation between genes influence the decrease in performance. For instance, while shuffling SHTN1 has the smallest impact on validation performance, its expression is significantly correlated with more genes than any other gene (with TPD52L1, NES and ST6GAL2, see Supplementary figure 3 for correlation matrices), and therefore losing its information in the signature is less impactful. Nevertheless, these results demonstrate that all individual genes are important for the validation performance, as none can be shuffled without decreasing performance.

**Figure 4.**
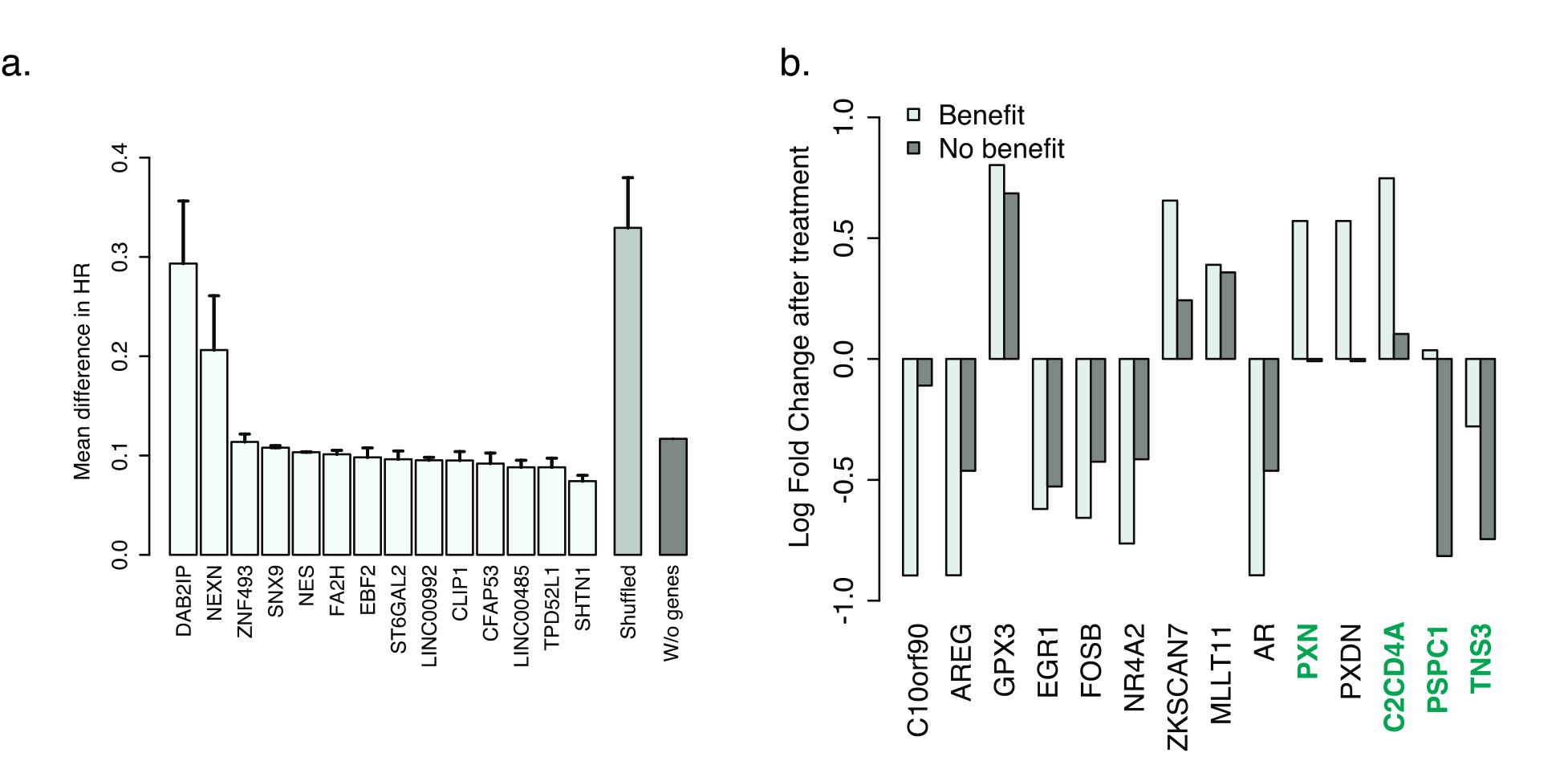
a. The decrease in performance (difference in HR) for i) shuffling of each gene separately, ii) shuffling links in the network and iii) when the 14 signature genes are excluded from the analysis. Error bars indicate standard error. b. Genes with a significant change in expression before and 48 hours after bortezomib treatment in only either class ‘benefit’ or ‘no benefit’. Genes in green have a significant difference in response between class ‘benefit’ and ‘no benefit’, determined empirically by testing the difference with 1000 random labellings.

Next, we assess whether the relation between the genes, as captured in the edges of the inferred network, are important for classifier performance. To this end, we shuffle the edges between all genes in the network, while ensuring every gene remains linked to at least one other gene, and then infer a signature with STLsig. This procedure is repeated 10 times. The mean HR found in the hold out data in class ‘benefit’ is 0.74 (se = 0.05), which is approximately equal to the HR found in the dataset without classification. This shows that, in addition to the individual genes, also their connections are essential for the performance of the signature.

Lastly, we remove all 14 genes contained in the signature from the dataset and search for a new signature by rerunning the entire STLsig approach. The new signature, which contains 312 genes from 85 gene networks, results in an HR of 0.56 (p = 0.23, 95% CI = 0.23 - 1.41) in the training data. This is a much worse HR than the original 14-gene signature. Also, the performance on the patients in the independent Fold D, which requires optimizing a new zPFS threshold, yields a worse performance (HR of 0.59; p = 0.06, 95% CI 0.34 - 1.01; n=130 in the ‘benefit’ class). Moreover, changing this threshold to yield a differently sized class ‘benefit’ does not yield performances that approach that of the original 14-gene signature (Supplementary Figure 1).

Together, these results establish that the 14 identified genes are essential to the performance of the model. This is an important prerequisite for interpretability of the model; as this indicates that the 14 genes may play a functional role in the proteasome inhibitor response.

### Multiple signature genes are associated with MM or proteasome inhibition

Having established the genes in the signature are essential to the performance, we investigate how the genes in the signature are involved in determining PI benefit. Interestingly, in addition to having the largest impact when its information is lost, DAB2IP is also the only gene that is significantly differentially expressed between class ‘benefit’ and ‘no benefit’ (p = 0.002). DAB2IP plays an essential role in the IRE1-mediated ER stress response and inducing apoptosis via the JNK pathway ^18^. Apoptosis induced by ER-stress is one of the main working mechanisms of proteasome inhibitors ^5^.

None of the other signature genes are significantly differentially expressed between the classes (Supplementary table 1). This lack of univariate association is to be expected, as our results show that the combination of genes is essential for the benefit prediction. Several genes do have a clear link to cancer or MM specifically. For instance, NES is a stem cell marker that is not found in healthy plasma cells, but is found specifically in MM^19^. Moreover, NES has been associated with treatment response in MM^19^. CLIP1 is involved in microtubule-kinetochore attachment and plays a role in proper chromosome alignment during mitosis ^19,20^ and has been associated with cancer progression and chemotherapy resistance ^21^, though not in relation to MM. SNX9 is described to play an important role in trafficking ADAM9 to the cell surface ^22^. ADAM9 is expressed in MM cells and induces IL6 production by osteoblasts, potentially creating a more permissive bone marrow environment for MM cell proliferation ^23^. One of the described working mechanisms of bortezomib is the downregulation of the production of IL-6 in the bone marrow environment ^23,24^. Some signature genes are also linked to the working of proteasome inhibitors. The gene TPD52L1 is a negative regulator of ATM ^25^, which is involved in the DNA damage response and activated by bortezomib treatment ^25,26^. ST6GAL2 has been described before to be significantly downregulated in carfilzomib-resistant cell lines ^27^.

Together, this indicates that our signature is not only capable of identifying patients benefiting from proteasome inhibitors, but also includes biologically relevant genes that aid in understanding differential response to PI treatment.

### Different cellular response to bortezomib in class benefit

For 142 patients in the HTT cohort tumor gene expression was measured again 48 hours after receiving bortezomib. To investigate whether the cellular response to bortezomib is different for patients classified as ‘benefit’, we performed a differential expression analysis between the gene expression before and after treatment separately in class ‘benefit’ and class ‘no benefit’ using SAM^28^. Because the number of patients in class ‘benefit’ for which a second measurement is available is small, we relaxed our definition of benefit and stratify the patients into the ‘benefit’ class if the calculated zPFS>0 and to the ‘no benefit’ class otherwise. This results in 71 out of 142 patients being classified as benefiting. We find 12 genes that are significantly differentially expressed before and after treatment in class ‘benefit’ but not in class ‘no benefit’. We also find two genes that are significantly differentially expressed only in class ‘no benefit’. These 14 differentially expressed genes and their mean fold change after treatment are shown in figure 4c. To identify the genes that truly represent a different cellular response in class ‘benefit’ and ‘no benefit’, we compute the difference in fold change between both classes. To ensure that this is not a random difference, we also compute this difference for all genes using 1000 random class labellings. We find four genes - TNS3, PXN, C2CD4A and PSPC1 - where the difference between ‘benefit’ and ‘no benefit’ is larger than expected by random chance (p <0.05 after Bonferroni correction for multiple testing). None of these genes have been described in the context of MM, though all have been connected to disease progression in other cancer types ^29,30,31,32^. More interestingly, TNS3, PXN and PSPC1 are all described to play a role in cell adhesion and a migratory phenotype ^33,32^. Cell adhesion mediated drug resistance (CAM-DR) has been described extensively in MM ^34,35,36^. Moreover, it has been suggested that bortezomib can overcome CAM-DR ^37,38^. A different regulation of cell adhesion in class ‘benefit’ could play a role in the observed benefit to PIs.

## Discussion

In this work we propose STLsig, a method to identify interpretable signatures that robustly predict patient benefit to PIs from a gene expression measurement at time of diagnosis. The 14 gene signature, derived with our method, validates on an independent patient cohort which was moreover measured on a different platform, confirming the robust nature of the signature.

PIs play a major role in MM treatment today. Yet our results indicate that many patients do not benefit from PI treatment and better selection of patients that do benefit is crucial. The signature identified here has direct applicability in the clinic to aid in optimal treatment selection.

One of the main aims of constructing simple gene expression signatures is that the genes in the signature may provide insight into the underlying biology. Interpreting the signature is, however, only meaningful if the genes in the signature are essential and unique. Our signature adheres to both these properties as substantial drop in performance is observed when one of the genes in the signature is shuffled and no similar signature can be found when the original 14 genes are excluded from the analysis. These findings reinforce the importance of the selected genes and indicate the power of STLsig to further elucidate proteasome inhibitor specific mechanisms.

Several of the genes in the signature are already described to be involved in the proteasome system or disease progression in MM. For instance, genes involved in the unfolded protein response have been described before to play a role in bortezomib response ^39^. The fact that our data-driven approach selects genes also involved in this process, indicates we can capture biology without inserting biological knowledge into the model beforehand. Therefore, we propose that the genes in our signature not yet described in the literature to be involved in PI efficacy, may have an as of yet undiscovered role in determining proteasome inhibitor benefit and are interesting targets for further research.

We found that our classifier, which was trained on bortezomib, also performed well in predicting benefit to another PI, carfilzomib. This is impressive as the model had no opportunity to learn carfilzomib-specific mechanisms. Although a carfilzomib signature that can be used in the clinic would need more specific optimization, our results do indicate that at least some sensitivity mechanisms overlap between bortezomib and carfilzomib.

STLsig can readily be applied to other diseases and drugs. A very potent application could be to perform post-hoc analysis of clinical trial data for drugs which missed their endpoint. Such analysis could reveal a subset of patients who would still benefit from the drug, thus potentially extracting extremely valuable information from failed clinical trials.

Taken together, we provide a powerful machine learning approach to aid in treatment decisions in the clinic, ensuring a more optimal treatment choice and ultimately improve patient outcomes.

## Methods

### Data

To develop the gene network and train the bortezomib benefit signature, we pool gene expression and survival data from three phase III trials (referred to as the HTT cohort): Total Therapy 2 (TT2, GSE2658)^40^, Total Therapy 3 (TT3, GSE2658)^40^ and HOVON-65/GMMG-HD4 (H65, GSE19784)^41^. The TT2 dataset includes 345 newly diagnosed multiple myeloma (NDMM) samples, treated either with thalidomide and melphalan (n = 173) or melphalan alone (n = 172). The TT3 dataset includes 238 NDMM samples treated with bortezomib, thalidomide, dexamethasone, cyclophosphamide, cisplatin and etoposide (VTDPACE). The H65 dataset includes 327 NDMM samples, treated either with vincristine, doxorubicin and dexamethasone (VAD, n = 158) or bortezomib, doxorubicin and dexamethasone (PAD, n = 169). In our analyses of the pooled data two treatment arms are considered: a bortezomib arm, which comprises the PAD arm from H65 and TT3, and a non-bortezomib arm, which comprises the VAD arm from H65 and TT2. Combined, these datasets form the HTT cohort and include 910 patients, of which 407 received bortezomib and 503 did not. Of these 910, 606 are used as HTT training data and 304 as HTT hold out data.

All samples have been profiled with the Affymetrix Human Genome U133 plus 2.0 array. Gene expression is MAS5 and log2 normalized. Batch effects resulting from pooling different datasets are corrected with ComBat ^42^. Data is scaled to mean 0 and variance 1 per probeset.

For validation of both the bortezomib model and carfilzomib model, we use the CoMMpass trial (NCT0145429) dataset generated by the Multiple Myeloma Research Foundation (MMRF). For 747 patients both RNAseq, survival data, and treatment information is available (CoMMpass Interim Analysis 13). Of these patients, 61 did not receive any PI in first line treatment, 530 received bortezomib and 156 received carfilzomib. Sequencing data is processed with the Cufflinks pipeline (for details see researcher.themmrf.org). For validation we combine the log2 normalized values from the HTT data and the FPKM values from CoMMpass. We scale the combined data to mean 0 and variance 1 and then perform ComBat batch correction, as performing mean-variance scaling before ComBat leads to better overlap between the datasets in the tSNE. In ComBat batch correction H65, TT2, TT3 and CoMMpass are defined as four separate batches.

For all survival analyses, we use Progression Free Survival (PFS) as an endpoint. Cox proportional hazard models were fitted using the R package ‘survival’ (version 2.44). All HRs are computed as PI vs no PI, which means a HR below 1 signifies a benefit from the treatment. For the survival analysis in the HTT cohort we stratify for dataset of origin, to correct for the significant survival difference between the Total Therapy dataset and the Hovon65/GMMG-HD 4 dataset.

### Constructing and evaluating gene pairs

We select only probe sets that meet the following requirements: (i) variance across the samples > 2 in the training dataset before mean variance scaling, (ii) unambiguous mapping to one gene and (iii) matching gene in the CoMMpass dataset. This yields n = 3319 genes. We then construct all possible gene pairs from these 3319 genes, resulting in 5,506,221 gene pairs.

To train the gene signature we divide the HTT cohort (n = 910) into four folds, Fold A (n = 202), Fold B (n = 202), Fold C (n = 202) and Fold D (n = 304). Fold A, B and C are used to train the signature as described below, while fold D acts as hold out data to validate the signature and optimize a threshold to use in independent validation data.

To determine treatment benefit, we follow the core concept of STL laid out in our previous work^3^, where for each patient a score zPFS is defined that measures whether the patient survived longer than expected compared to patients with similar gene expression that received another treatment. More specifically, for genepair {n,m} and patient j we define:

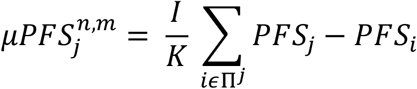

where PFS_j_ is the progression free survival time of patient *j*, I=1 if patient j received the target treatment (a PI in this work) and I=-1, otherwise. Moreover, Π^*j*^ the set of K nearest neighbors to patient j defined in terms of euclidean distance in the expression space spanned by genes n and m and only considering patients that received another treatment than patient j. Throughout this manuscript K=10. In the set Π^*j*^, we discard patients for whom we cannot be sure whether they survived longer or not (i.e. if both patients are censored). This leads to an average of 7 patients being used in the calculation of μPFS_j_. Subsequently, zPFS is normalized to a z-score by comparing *μ*PFS to a background distribution resulting from repeating this procedure M=1000 times with a random Π^*j*^. The zPFS score describes how much smaller (or larger) the survival of patient j is compared to patients with similar gene expression but opposite treatment than expected by random chance.

To score gene pairs, a 2-fold cross validation is employed using fold A (n = 202) and fold B (n = 202). Within each fold, a kNN-regression model (k = 10) is trained, which is used to predict zPFS on the other fold. The gene pair score is defined as the Spearman correlation coefficient between the predicted zPFS and calculated zPFS across all patients. The score for each gene pair is the mean correlation of the 2 folds. We repeat this procedure 5 times with a different split in folds.

### Gene network construction

We construct gene networks separately for all 5 repeats and then construct a consensus network, which only contains the genes and edges found in all 5 repeats. To construct the gene networks, for each gene, we rank all gene pairs containing that gene on the mean Spearman correlation coefficient found. We then connect genes that are mutually synergistic, i.e. gene A should be among gene B’s best partners and vice versa. We achieve this by requiring that AB is among the top 5% of pairs including A and among the top 5% of pairs including B. However, if a single gene is informative for treatment benefit, gene pairs containing this gene could be highly ranked even if the second gene is uninformative. Including these gene pairs in our network and subsequent signature would introduce noise, which would both harm biological interpretation of the signature and potentially decrease the predictive performance in independent data. Therefore, we also require the mean correlation of the gene pair to be above the median correlation of all selected gene pairs. We evaluate all gene networks in the consensus network on their ability to predict benefit and select the best performing combination to construct the signature.

### Gene network selection and gene signature construction

After gene network construction, gene networks are selected using forward feature selection. To rank gene networks, we determine the predictive performance for each gene network. To this end, we calculate zPFS for each patient and each gene network separately on fold A and B together (n = 404). The top 25% of patients (in terms of zPFS) are assigned to class ‘benefit’, while the remaining patients are assigned to class ‘no benefit’. Subsequently, A Cox proportional hazards regression on the treatment variable is performed within the ‘benefit’ patient group as well as in the ‘no benefit’ patients. The performance of a gene network is defined by the difference between the Cox’ regression β’s in class ‘benefit’ and class ‘no benefit’.

To select gene networks to use in the final model we perform forward feature selection using fold C, which comprises 202 patients not used in fold A and B. Gene networks are added sequentially based on their performance on fold A and B. Ranking of patients across more than one gene network is done based on the sum of the zPFS scores of the individual gene networks.

### Validation of gene networks

To validate the signature in independent data, we use all training data (n = 606) as a reference set where zPFS is known. For each patient in the validation set we compute the euclidean distance to all patients in the reference set per gene network. We then use inverse distance weighting to calculate the estimated zPFS of a validation patient *j* by

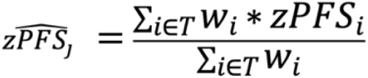

where *T* comprises all patients in the reference dataset. Given a certain gene expression vector ***x***, weights *w*_*i*_ are given by

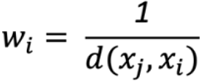

where d is the Euclidean distance between the expression data of gene x of patients i and j.

## Data Availability

The datasets supporting the conclusions of this article are available on GEO. Gene expression data from the HOVON-65/GMMG-HD4 study is available at GSE19784 (https://www.ncbi.nlm.nih.gov/geo/query/acc.cgi?acc=GSE19784). Gene expression data from both Total Therapy 2 and Total Therapy 3 are available at GSE2658 (https://www.ncbi.nlm.nih.gov/geo/query/acc.cgi?acc=GSE2658). 30 patients from the Total Therapy 3 study used in the manuscript are not included in the GSE2658 dataset, these can be found in ArrayExpress dataset E-TABM-1138 (https://www.ebi.ac.uk/arrayexpress/experiments/E-TABM-1138/). The PFS survival data for all three studies are available at https://github.com/jubels/GESTURE, linked to the GEO and ArrayExpress IDs. All gene expression and survival data for the CoMMpass study is available at research.themmrf.org

https://www.ncbi.nlm.nih.gov/geo/query/acc.cgi?acc=GSE2658

https://www.ebi.ac.uk/arrayexpress/experiments/E-TABM-1138/

https://research.themmrf.org

## Declarations

### Ethics approval

Not applicable

### Consent for publication

Not applicable

### Data and code availability

All code needed to discover and validate the signature is available at https://github.com/jubels/STLsig. All code requires R and is platform independent.

### Author contributions

J.d.R., M.H.v.V., and J.U. developed the algorithm. J.U. implemented the algorithm and performed data analysis. J.d.R. and J.U. wrote the manuscript. M.H.v.V and P.S. provided comments and edits. All authors discussed the results.

## Acknowledgements

The CoMMpass dataset was generated as part of the Multiple Myeloma Research Foundation Personalized Medicine Initiatives (https://research.themmrf.org and www.themmrf.org).

## Figure legends

**Supplementary figure 1.**
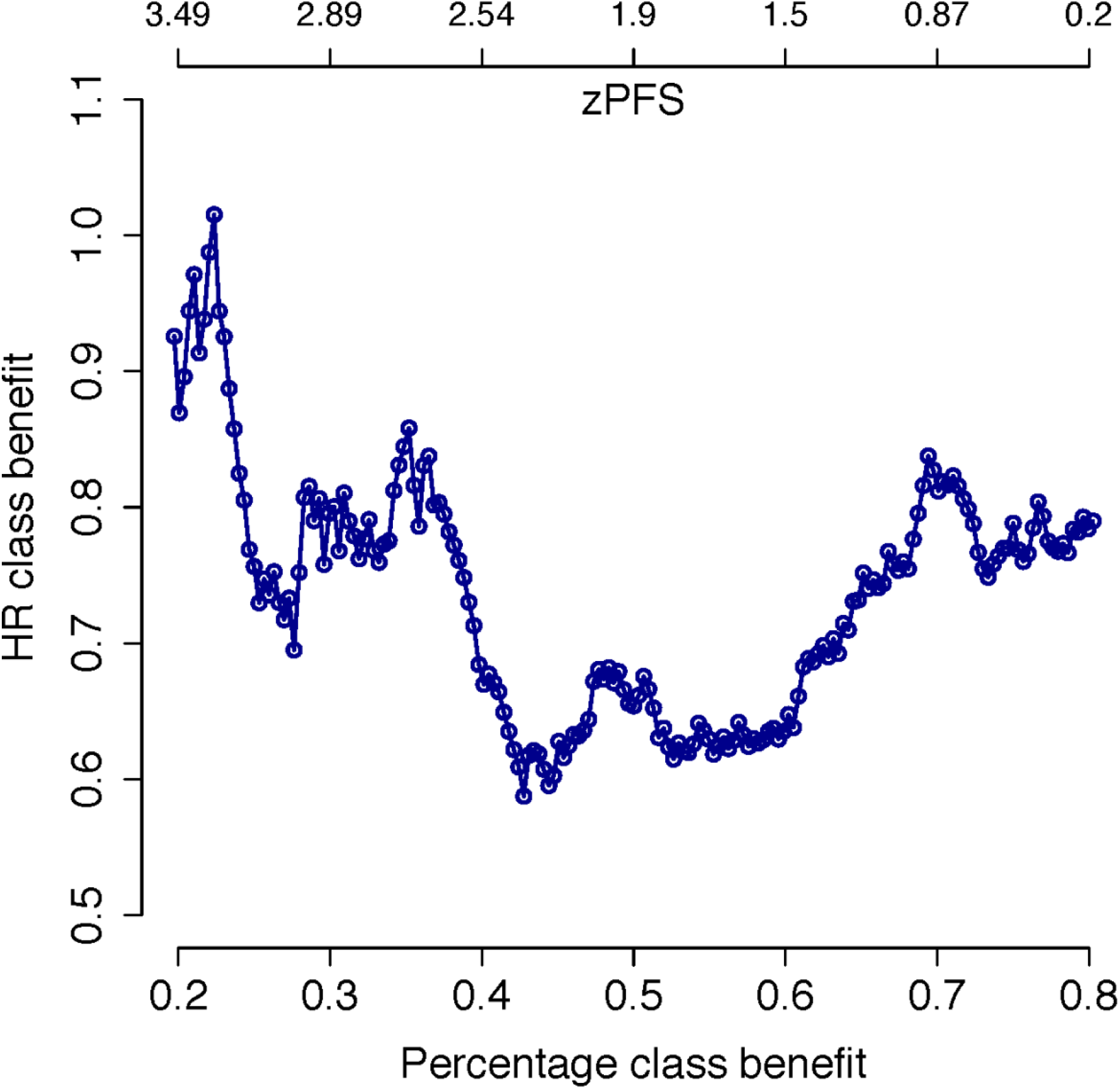
The HR found in class ‘benefit’ using different cutoffs for zPFS, as predicted by the signature found when excluding the original 14 genes from the analysis.

**Supplementary figure 2.**
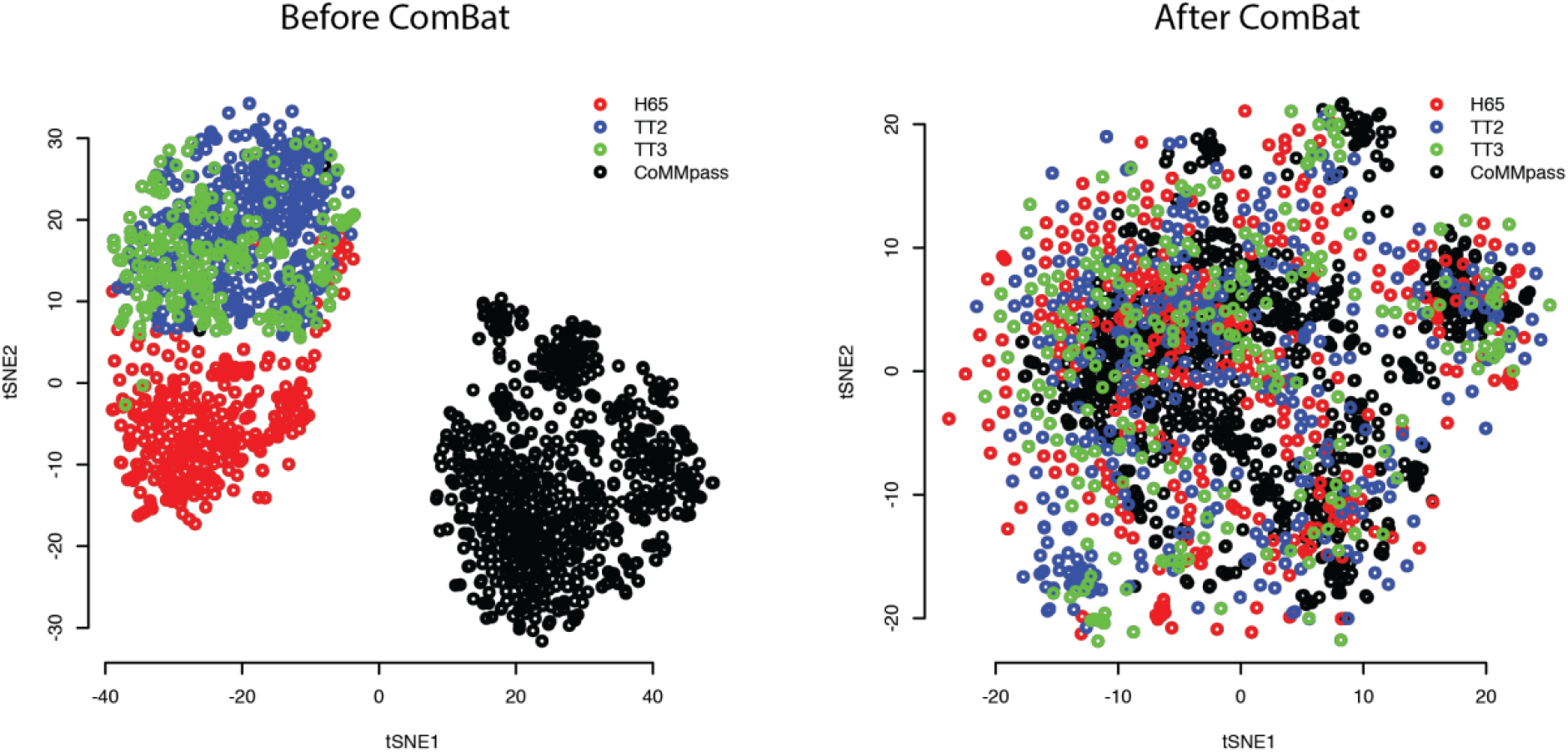
tSNE of the datasets before and after batch correction with ComBat

**Supplementary figure 3.**
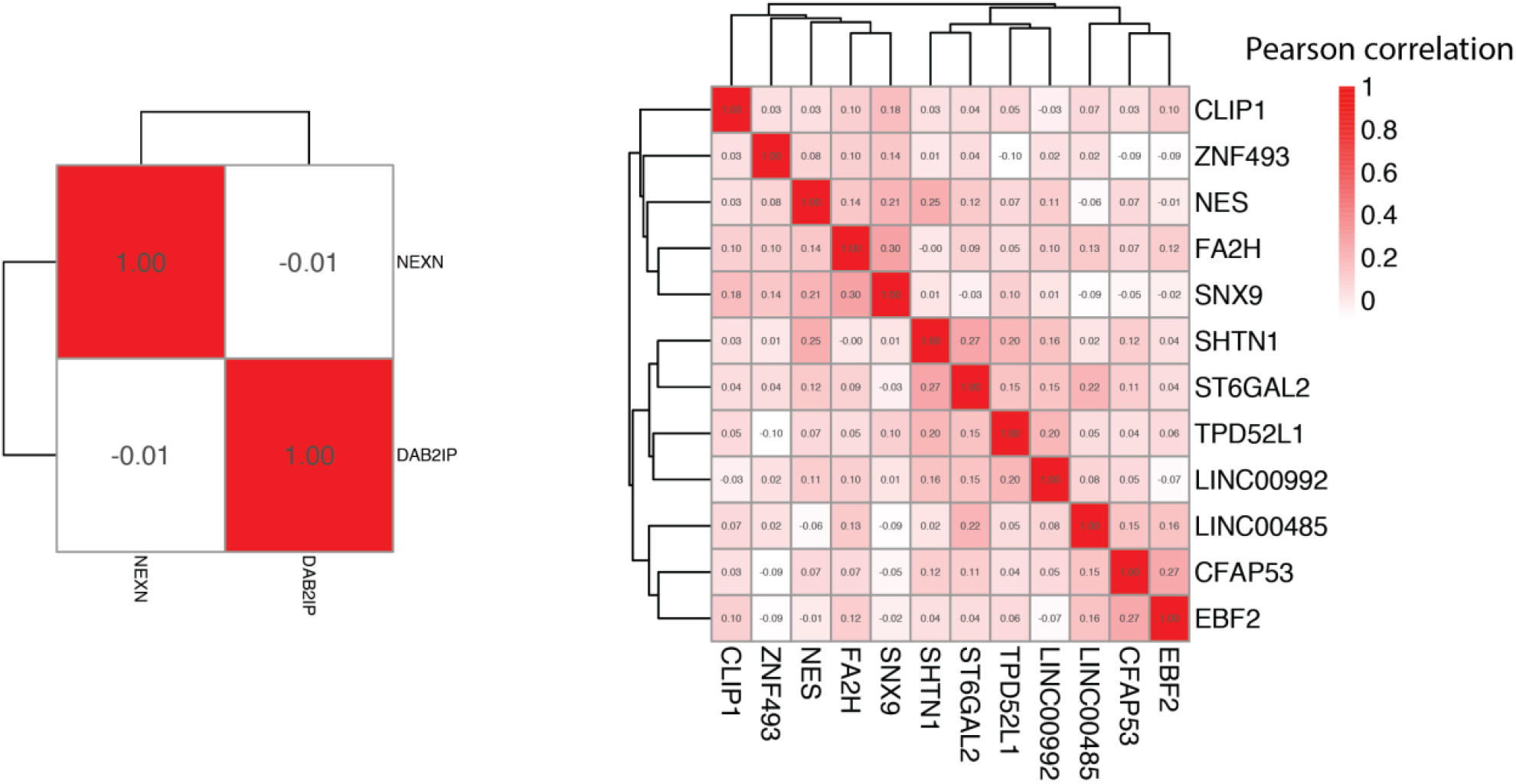
Correlation matrix of the Pearson correlation between the gene expression of the genes included in the signature, per gene network.

**Supplementary table 1.** Log2 fold difference of signature genes between class ‘benefit’ and ‘no benefit’ in fold D of the HTT cohort

|  | Mean log2 fold difference | p-value |
| --- | --- | --- |
| NEXN | 0.27 | 0.16 |
| DAB2IP | -0.65 | 0.002 |
| CFAP53 | -0.06 | 0.72 |
| TPD52L1 | 0.20 | 0.34 |
| SHTN1 | 0.38 | 0.05 |
| ZNF493 | -0.015 | 0.95 |
| NES | 0.10 | 0.77 |
| CLIP1 | 0.23 | 0.26 |
| LINC0048<br>5 | 0.15 | 0.49 |
| ST6GAL2 | 0.26 | 0.14 |
| EBF2 | -0.14 | 0.44 |
| LINC0099<br>2 | 0.15 | 0.44 |
| FA2H | 0.05 | 0.74 |
| SNX9 | -0.006 | 0.98 |

